# Role of the repeat expansion size in predicting age of onset and severity in RFC1 disease

**DOI:** 10.1101/2023.10.15.23297048

**Authors:** Riccardo Currò, Natalia Dominik, Stefano Facchini, Elisa Vegezzi, Roisin Sullivan, Valentina Galassi Deforie, Gorka Fernández-Eulate, Andreas Traschütz, Salvatore Rossi, Matteo Garibaldi, Mariusz Kwarciany, Franco Taroni, Alfredo Brusco, Jean-Marc Good, Francesca Cavalcanti, Simon Hammans, Gianina Ravenscroft, Richard H Roxburgh, RFC1 repeat expansion study group, Ricardo Parolin Schnekenberg, Bianca Rugginini, Elena Abati, Arianna Manini, Ilaria Quartesan, Arianna Ghia, Adolfo Lòpez de Munaìn, Fiore Manganelli, Marina Kennerson, Filippo Maria Santorelli, Jon Infante, Wilson Marques, Manu Jokela, Sinéad M Murphy, Paola Mandich, Gian Maria Fabrizi, Chiara Briani, David Gosal, Davide Pareyson, Alberto Ferrari, Ferran Prados, Tarek Yousry, Vikram Khurana, Sheng-Han Kuo, James Miller, Claire Troakes, Zane Jaunmuktane, Paola Giunti, Annette Hartmann, Nazli Basak, Matthis Synofzik, Tanya Stojkovic, Marios Hadjivassiliou, Mary M Reilly, Henry Houlden, Andrea Cortese

## Abstract

*RFC1* disease, caused by biallelic repeat expansion in *RFC1*, is clinically heterogeneous in terms of age of onset, disease progression and phenotype. We investigated the role of the repeat size in influencing clinical variables in *RFC1* disease. We also assessed the presence and role of meiotic and somatic instability of the repeat.

In this study, we identified 553 patients carrying biallelic *RFC1* expansions and measured the repeat expansion size in 392 cases. Pearson’s coefficient was calculated to assess the correlation between the repeat size and age at disease onset. A Cox model with robust cluster standard errors was adopted to describe the effect of repeat size on age at disease onset, on age at onset of each individual symptoms, and on disease progression. A quasi-poisson regression model was used to analyse the relationship between phenotype and repeat size. We performed multi-variate linear regression to assess the association of the repeat size with the degree of cerebellar atrophy. Meiotic stability was assessed by Southern blotting on first-degree relatives of 27 probands. Finally, somatic instability was investigated by optical genome mapping on cerebellar and frontal cortex and unaffected peripheral tissue from four post-mortem cases.

A larger repeat size of both smaller and larger allele was associated with an earlier age at neurological onset (smaller allele HR=2.06, p<0.001; larger allele HR=1.53, p<0.001) and with a higher hazard of developing disabling symptoms, such as dysarthria or dysphagia (smaller allele HR=3.40, p<0.001; larger allele HR=1.71, p=0.002) or loss of independent walking (smaller allele HR=2.78, p<0.001; larger allele HR=1.60; p<0.001) earlier in disease course. Patients with more complex phenotypes carried larger expansions (smaller allele: complex neuropathy RR=1.30, p=0.003; CANVAS RR=1.34, p<0.001; larger allele: complex neuropathy RR=1.33, p=0.008; CANVAS RR=1.31, p=0.009). Furthermore, larger repeat expansions in the smaller allele were associated with more pronounced cerebellar vermis atrophy (lobules I-V β=-1.06, p<0.001; lobules VI-VII β=-0.34, p=0.005). The repeat did not show significant instability during vertical transmission and across different tissues and brain regions.

*RFC1* repeat size, particularly of the smaller allele, is one of the determinants of variability in *RFC1* disease and represents a key prognostic factor to predict disease onset, phenotype, and severity. Assessing the repeat size is warranted as part of the diagnostic test for *RFC1* expansion.

## Introduction

Repeat expansion disorders are a group of diseases caused by abnormally long microsatellites, also called short tandem repeats, located either in coding or non-coding regions of the human genome.^1–4^ The recent developments in whole-genome sequencing methods have led to an increased identification and diagnosis of diseases caused by non-coding repeat expansion.^5,6^ Short tandem repeats are dynamic elements that are variably prone to further expand in offspring and across different tissues from the same individual, leading to genetic anticipation and, arguably, selective tissue involvement.^7–13^ Notably, repeat expansion disorders typically show a correlation between the repeat length and an earlier onset and more severe disease phenotype.^10,14–22^

Biallelic expansion of AAGGG pentanucleotides *(TTCCC in the transcription sense)* in the second intron of the replication factor complex subunit 1 (*RFC1*) was identified as the main cause of cerebellar ataxia, neuropathy and vestibular areflexia syndrome (CANVAS)^23,24^ and subsequent studies reported a high prevalence of biallelic AAGGG expansions in cases with sporadic or familial ataxia.^23,25–29^ To date, biallelic AAGGG expansions explain the vast majority of CANVAS cases, with only few cases recently reported to carry different, population-specific configurations (that is, ACAGG or AAAGG_10–25_AAGGG_exp_AAAGG_4–6_ in East Asia and Oceania^28,30,31^) or mono-allelic AAGGG expansions in compound heterozygous state with truncating variants.^32–35^ A sensory neuropathy was recognized as the key feature in *RFC1* disease spectrum and up to a third of cases diagnosed with idiopathic sensory neuropathy, with or without ataxia and vestibular impairment, carry biallelic *RFC1* expansions.^36,37^ Clinical heterogeneity in *RFC1* disease also involves the disease course and severity, as revealed by the wide range of age at onset and disability.^29,36,38^ However, determinants of the variability of *RFC1* disease are still largely unknown.

In this multicentre study we assessed the impact of the AAGGG repeat expansion size on the disease onset, phenotype, and severity in a large cohort of patients carrying biallelic *RFC1* expansions. To gain further insight into the intergenerational transmission and disease pathogenesis, we also investigated the stability of the AAGGG repeat in families and within different tissues of affected individuals.

## Materials and methods

### Patients

The study population consisted of a multicentre cohort of 2334 patients diagnosed with sensory neuropathy, adult-onset (>25 years old) cerebellar ataxia, complex neuropathy, or CANVAS. Sensory neuropathy was diagnosed according to clinical and neurophysiological criteria.^39,40^ Complex neuropathy was defined by the presence of sensory neuropathy and evidence of either cerebellar or vestibular involvement on examination and/or investigations. Patients with combined involvement of sensory, vestibular and cerebellar systems were classified as CANVAS.^41,42^ Previous studies demonstrated that sensory involvement is the hallmark of *RFC1* disorder^36,38^. Accordingly, a phenotype category was not assigned to patients whose sensory examination or nerve conduction studies were not available. Furthermore, clinical phenotype was defined only when at least two of the three core systems (i.e., sensory, cerebellar, and vestibular system) were examined.

### Clinical features

Clinical and demographic data of patients with positive genetic testing for biallelic *RFC1* expansions were collected according to a standardised template which was completed by all referring clinicians and which included: family history, age at onset of any neurological symptom, including sensory symptoms, dysarthria, dysphagia, and oscillopsia, use of walking aids, and detailed first and last available neurological examinations. To avoid a possible confounder effect of population-specific non canonical configurations^28,30,31^, we included only patients of Caucasian ancestry in our analyses. The presence of chronic cough was also recorded, but it was not considered to define the neurological onset of the condition. Assessment of sensory system was available in 381 cases (97%), of cerebellum in 385 (98%) and of vestibular system in 260 (66%). Based on the presence of symptoms and signs, patients were divided into three categories: sensory neuropathy, complex neuropathy, and CANVAS. Additional features, such as parkinsonism, cognitive impairment, symptomatic dysautonomia or pyramidal involvement were also recorded. Dysarthria and dysphagia and loss of independent walking were further analysed as markers of disease severity.

### Brain MRI data acquisition

Structural T1 magnetic resonance images of 59 brain MRI acquired from 2004 to 2023 in a clinical setting at the National Hospital for Neurology and Neurosurgery (London, UK) were retrieved for volumetric analyses. 2D or 3D acquisitions were included depending on availability. Twenty-seven MRI were discarded as they did not pass quality check. Brain parcellations were computed using Geodesic Information Flows (GIF)^43^ (GIF is free and available as webservice in NityWeb^44^) and following the Desikan-Killiany-Tourville atlas^45^. After parcellation, volumes were separately computed for cerebellar vermal lobules I-V, lobules VI-VII, lobules VII-X, and for the total intracranial volume (TIV) in mm^3^.

### PCR-based screening of RFC1 AAGGG repeat expansion

*RFC1* genetic test was performed as previously described.^23,42^ Samples with no amplifiable products on flanking PCR and positive repeat-primed PCR (RP-PCR) for the AAGGG repeat were considered likely positive for biallelic AAGGG *RFC1* expansions, after the exclusion of the non-pathogenic AAAGG and AAAAG expansions on the other allele.

### Southern blotting

Provided that enough DNA with good quality was available, samples were analysed by Southern Blotting as previously described,^23^ to confirm the presence and to measure the size of the expanded alleles. The lowest size for AAGGG repeat expansions detected in this study was 6.5 kilobases (that is approximately 250 repeat units).

### Meiotic instability

DNA from affected or unaffected first-degree relatives of index cases from 27 families was tested by Southern Blotting. *RFC1* repeat size within families was compared to evaluate the stability of the AAGGG repeat during intergenerational transmission.

### Somatic instability

Optical genome mapping (OGM; Bionano Genomics, San Diego, USA) was performed to assess the presence of post-zygotic instability in affected (vermis, cerebellar hemispheres) versus unaffected tissues (frontal cortex, muscle, fibroblasts) of patients carrying biallelic *RFC1* expansions. OGM has shown a good correlation with Southern Blotting in the identification and sizing of large repeat expansions, including *RFC1*,^46^ and a higher sensitivity in detecting the presence of somatic variation.^47^ Blood-derived DNA from a patient with *C9orf72* GGGGCC expansion was also included as positive control for repeat instability.^48^ Samples were processed as previously described^47^. Labelled ultra-high molecular weight (UHMW) gDNA was loaded on a Saphyr chip for linearization and imaging on the Saphyr instrument (Bionano Genomics, San Diego, CA, USA). The repeat expansion size was estimated as the difference between the mean of the Gaussian distribution of molecules mapping to the expanded alleles and the reference intermarker distance. Repeat sizes in different tissues and their standard deviations were calculated and compared to detect somatic instability.

### Statistical analysis

Data were expressed as means with standard deviations or medians with 25%-75% interquartile ranges (IQRs) and min-max values depending on their distribution. Statistical significance threshold was set to p<0.05 and correction for multiple comparisons was applied, as appropriate. We have accounted for the presence of clustered data (i.e., members of the same families) adopting cluster-adjusted robust standard errors in survival models and by adding a family random effect in linear regressions. To address the problem of collinearity due to the correlation between the repeat size of smaller and larger allele, all the analyses were performed adopting two separate models, one for each allele. First, we calculated Pearson’s correlation coefficients for repeat size of the smaller and larger allele and age at disease onset (cough excluded). We then ran a Cox regression to evaluate the effect of repeat size on age at disease onset (cough excluded) and at onset of main disease symptoms (i.e., unsteadiness, sensory symptoms, dysarthria and/or dysphagia, oscillopsia, chronic cough). Time from disease onset to dysarthria and /or dysphagia and to use of walking aids were considered as outcomes to predict the effect of repeat size on progression to disabling disease. A Fine-Gray competing risk model was adopted to adjust for competing risk (i.e., risk of the patient dying before experiencing the symptom). For each regression model, regression tables with Hazard Ratios (HR), 95% confidence intervals (CI), and p-value of a two-tailed Wald’s test on the coefficients for a 1000-unit change in repeat size were reported (*supplementary materials*). Predicted Cumulative Incidence Functions (CIF) were plotted for all the symptoms of interest. A quasi-poisson regression model was used to analyse the relationship between phenotype and number of repeat units. Coefficients were reported as Rate Ratios (RR). The model was adjusted for sex, age at last examination and disease duration, and was followed by Tukey adjusted pairwise comparisons between the three phenotypes. Multivariate linear regression was performed to assess the correlation between the repeat size and the degree of atrophy of cerebellar vermis, adjusted for age, disease duration, and total intraparenchymal volume (TIV). Meiotic stability of the repeat was assessed by a linear mixed-effects model. All analyses were performed using STATA statistical software, version 14. Plots and graphs were created with GraphPad Prism version 9.4.1 for Windows, GraphPad Software, San Diego, California USA, www.graphpad.com.

### Ethics

The study was approved by the ethics committee and by local institutional review boards. All patients gave informed consent prior to their inclusion in the study. The study complied with all relevant ethical regulations.

## Results

### Genetic testing for RFC1 expansions

Out of 2334 patients, 556 (24%) carried biallelic AAGGG expansions at PCR screening. A sufficient amount of good quality DNA to perform Southern Blotting was available in 395 cases. We confirmed the biallelic expansions in 392 patients (99.3%). Sanger sequencing in the three unconfirmed samples showed intermediate expansions (<100 repeats) of non-pathogenic AAAAG, AAAGG or AAAGGG motifs, which were missed by the previous PCR-based screening (**figure 1**).

**Figure 1.**
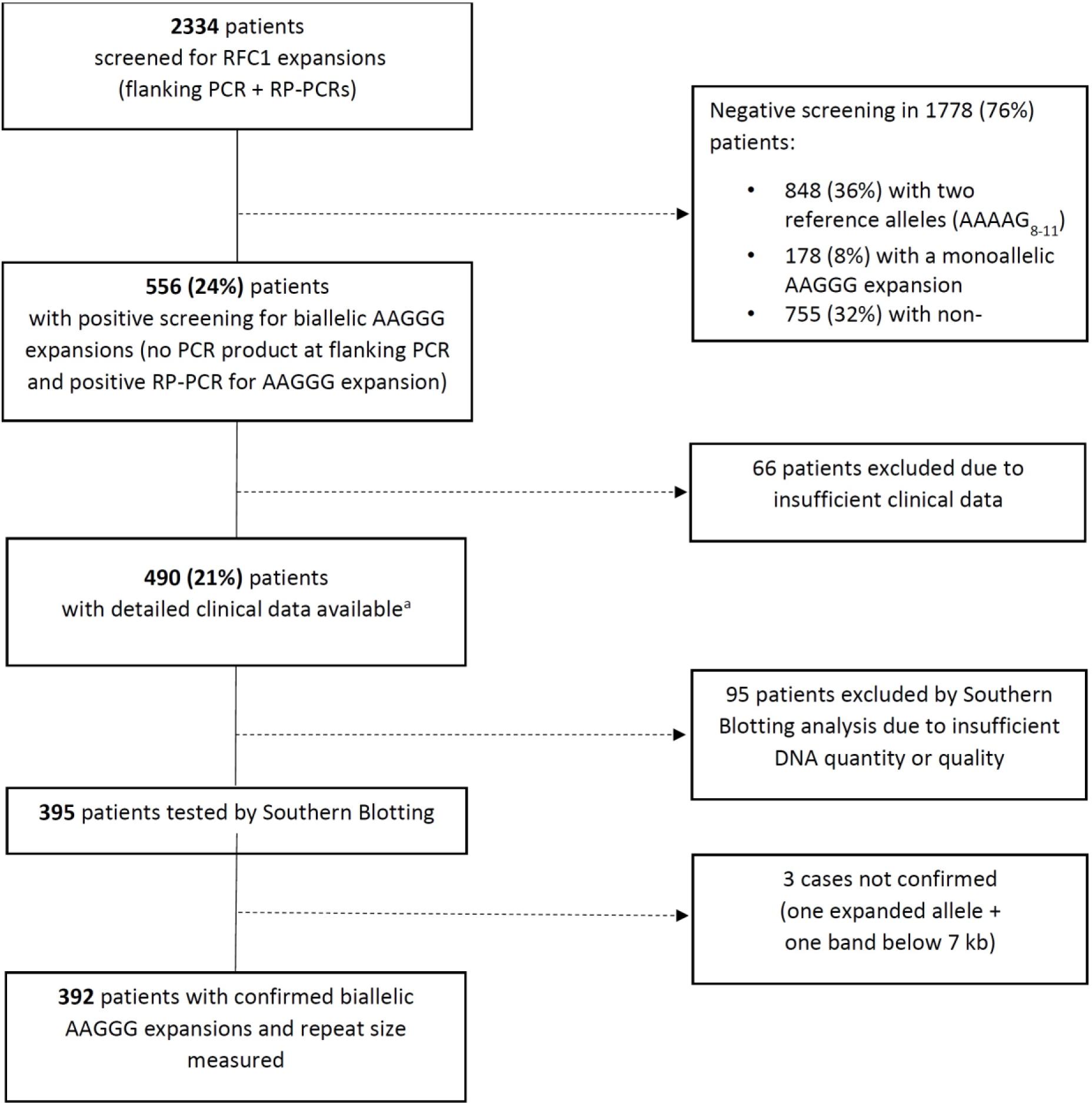
Flowchart describing the results of the genetic screening for RFC1 expansions. PCR= polymerase chain reaction; RP-PCR= repeat-primed polymerase chain reaction, kb= kilobases. ^a^Three cases were subsequently excluded from the analysis because Southern Blotting did not confirm the presence of biallelic expansions.

### Clinical heterogeneity and disease course of RFC1 disease

Demographic and clinical data from the 392 patients confirmed at Southern Blotting are summarised in **table 1**. There was a similar number of males and females. Three hundred and forty-seven cases were sporadic (89%), 45 cases were familiar from 19 families, including 14 families with 2 members, 3 with 3 members, and 2 families with 4 members affected. All cases were Caucasian and most of them (*n*=363, 92%) from European descents, however multiple countries were represented, including Turkey (*n*=18), Brazil (*n*=1), Iran (*n*=1), Iraq (*n*=1), Algeria (*n*=1), Lebanon (*n*=1). Country of origin was not specified for 6 patients. Median age at onset of neurological symptoms (cough excluded) was 54 years (*IQR*=49-61), ranging from 25 to 80 years. Unsteadiness was the most common complaint at disease onset, followed by sensory symptoms (e.g., loss of feeling, tingling, pins-and-needles). Dysarthria and/or dysphagia, suggestive of cerebellar involvement, and oscillopsia, due to bilateral vestibular impairment, were less frequent in the initial stages of disease but were present in up to 51% and 27% of patients at the most recent evaluation, respectively. Chronic cough was investigated in 358 patients (91%) and reported by 267 of them (75%). Cough was the presenting symptom in half of the cases. Fifty-four per cent of patients required walking aids after a median disease duration of 10 years (*IQR*=5-16) and 17% needed a wheelchair after 14 years (*IQR=*11-21). Less common symptoms and signs included: symptomatic dysautonomia (*n*=17), cognitive impairment (*n*=7), parkinsonism (*n*=2), upper motor neuron involvement other than brisk reflexes (e.g., spasticity, Babinski sign) (*n*=1).

**Table 1:**
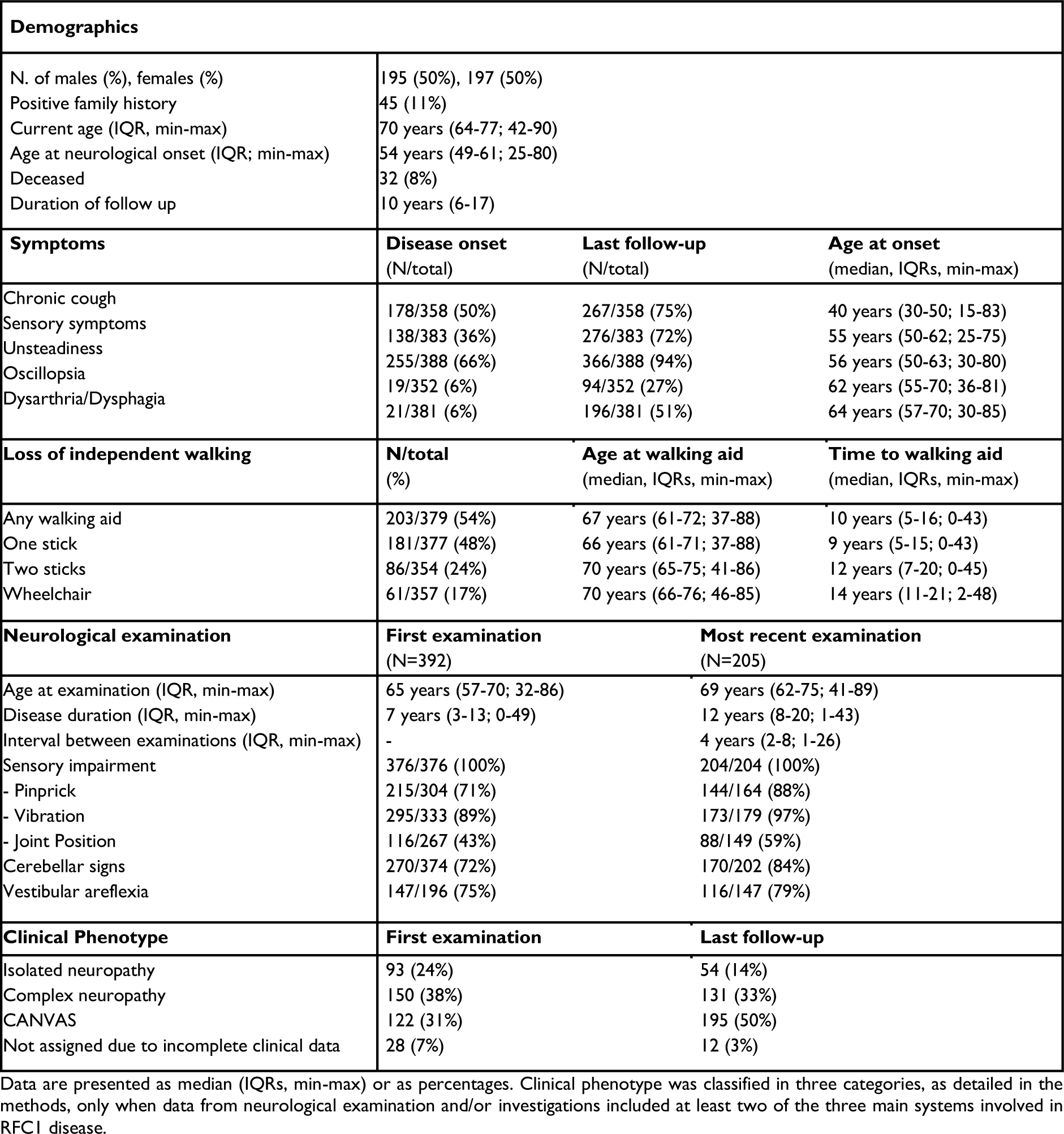
Demographic and clinical data of RFC1 positive patients.

| Demographics |  |  |  |
| --- | --- | --- | --- |
| N. of males (%), females (%) | 195 (50%), 197 (50%) |  |  |
| Positive family history | 45 (11%) |  |  |
| Current age (IQR, min-max) | 70 years (64-77; 42-90) |  |  |
| Age at neurological onset (IQR; min-max) | 54 years (49-61; 25-80) |  |  |
| Deceased | 32 (8%) |  |  |
| Duration of follow up | 10 years (6-17) |  |  |
| Symptoms | Disease onset<br>(N/total) | Last follow-up<br>(N/total) | Age at onset<br>(median, IQRs, min-max) |
| Chronic cough | 178/358 (50%) | 267/358 (75%) | 40 years (30-50; 15-83) |
| Sensory symptoms | 138/383 (36%) | 276/383 (72%) | 55 years (50-62; 25-75) |
| Unsteadiness | 255/388 (66%) | 366/388 (94%) | 56 years (50-63; 30-80) |
| Oscillopsia | 19/352 (6%) | 94/352 (27%) | 62 years (55-70; 36-81) |
| Dysarthria/Dysphagia | 21/381 (6%) | 196/381 (51%) | 64 years (57-70; 30-85) |
| Loss of independent walking | N/total<br>(%) | Age at walking aid<br>(median, IQRs, min-max) | Time to walking aid<br>(median, IQRs, min-max) |
| Any walking aid | 203/379 (54%) | 67 years (61-72; 37-88) | 10 years (5-16; 0-43) |
| One stick | 181/377 (48%) | 66 years (61-71; 37-88) | 9 years (5-15; 0-43) |
| Two sticks | 86/354 (24%) | 70 years (65-75; 41-86) | 12 years (7-20; 0-45) |
| Wheelchair | 61/357 (17%) | 70 years (66-76; 46-85) | 14 years (11-21; 2-48) |
| Neurological examination | First examination<br>(N=392) | Most recent examination<br>(N=205) |  |
| Age at examination (IQR, min-max) | 65 years (57-70; 32-86) | 69 years (62-75; 41-89) |  |
| Disease duration (IQR, min-max) | 7 years (3-13; 0-49) | 12 years (8-20; 1-43) |  |
| Interval between examinations (IQR, min-max) | - | 4 years (2-8; 1-26) |  |
| Sensory impairment | 376/376 (100%) | 204/204 (100%) |  |
| - Pinprick | 215/304 (71%) | 144/164 (88%) |  |
| - Vibration | 295/333 (89%) | 173/179 (97%) |  |
| - Joint Position | 116/267 (43%) | 88/149 (59%) |  |
| Cerebellar signs | 270/374 (72%) | 170/202 (84%) |  |
| Vestibular areflexia | 147/196 (75%) | 116/147 (79%) |  |
| Clinical Phenotype | First examination | Last follow-up |  |
| Isolated neuropathy | 93 (24%) | 54 (14%) |  |
| Complex neuropathy | 150 (38%) | 131 (33%) |  |
| CANVAS | 122 (31%) | 195 (50%) |  |
| Not assigned due to incomplete clinical data | 28 (7%) | 12 (3%) |  |

Data are presented as median (IQRs, min-max) or as percentages. Clinical phenotype was classified in three categories, as detailed in the methods, only when data from neurological examination and/or investigations included at least two of the three main systems involved in RFC1 disease.

Patients had a first neurological assessment at a median age of 65 years (*IQR*=57-70), after a median disease duration of 7 years (*IQR*=3-13). All cases had signs and/or symptoms of sensory neuropathy, when investigated. Ninety-three patients (24%) had an isolated sensory neuropathy, 150 (38%) a complex neuropathy with vestibular or cerebellar involvement and 122 (31%) CANVAS. A phenotype was not assigned in 28 cases (7%) due to incomplete clinical assessment. A second examination was available in 205 cases, after a median interval of 4 years (*IQR*=2-8) from the first examination and of 12 years (*IQR*=8-20) from disease onset. Additional 12% and 17% patients developed signs of cerebellar and vestibular involvement, respectively. Overall, 195 patients (50%) had complete CANVAS, 131 (33%) had a complex neuropathy, while 54 (14%) still showed an isolated sensory neuropathy.

Thirty-two patients (8%) were deceased at the time of the study and in 7 cases death was related to the underlying neurological disease (i.e., four due to complications of prolonged immobility or falls, three due to aspiration pneumonia or cachexia in dysphagic patients). Other reported causes of death were neoplasms (*n*=3), Sars-Cov2 infection (*n*=2), myocardial infarction (*n*=1), cerebral haemorrhage (*n*=1), pulmonary fibrosis and respiratory failure (*n*=2). Median age at death was 76 years (*IQR=*74-78) for men and 76 years (*IQR*=74-79) for women, which is slightly below the mean life expectancy in Europe according to WHO data (overall=80.4; females=83.2; males=77.5).^49^

### Repeat expansion size predicts onset and progression of RFC1 disease

The median number of AAGGG repeats was 1042 (*IQR=*844-1306; *range*=249-3885), and specifically 937 repeat units (*IQR=*771-1129; *range=*249-2411) for the smaller allele and 1195 repeat units (*IQR=*927-1452; *range=*294-3885) for the larger allele. Notably, we observed a significant correlation (*r*=0.7, *p*<0.001) between the size of the two alleles. In 143 patients (36%) the two expanded alleles appeared as a single band on Southern Blotting. This suggests that they had the same or highly similar size, within the limits of detection resolution of this technique.

Detailed tables and figures for the statistical analyses are provided in the supplementary materials. We observed an inverse correlation between age at neurological onset and repeat size of the smaller allele (*r*=-0.21, *r^2^*=0.06, *p*<0.001) and the larger allele (*r*=-0.17, *r^2^*=0.03, *p*<0.001) (**figure 2A-2B**). After adjusting for sex and clinical phenotype, the association with age at neurological onset was still more significant for the smaller allele (*HR*=2.06, *p*<0.001) than for the larger allele (*HR*=1.53, *p*<0.001) **(supplementary table 1s).**

**Figure 2.**
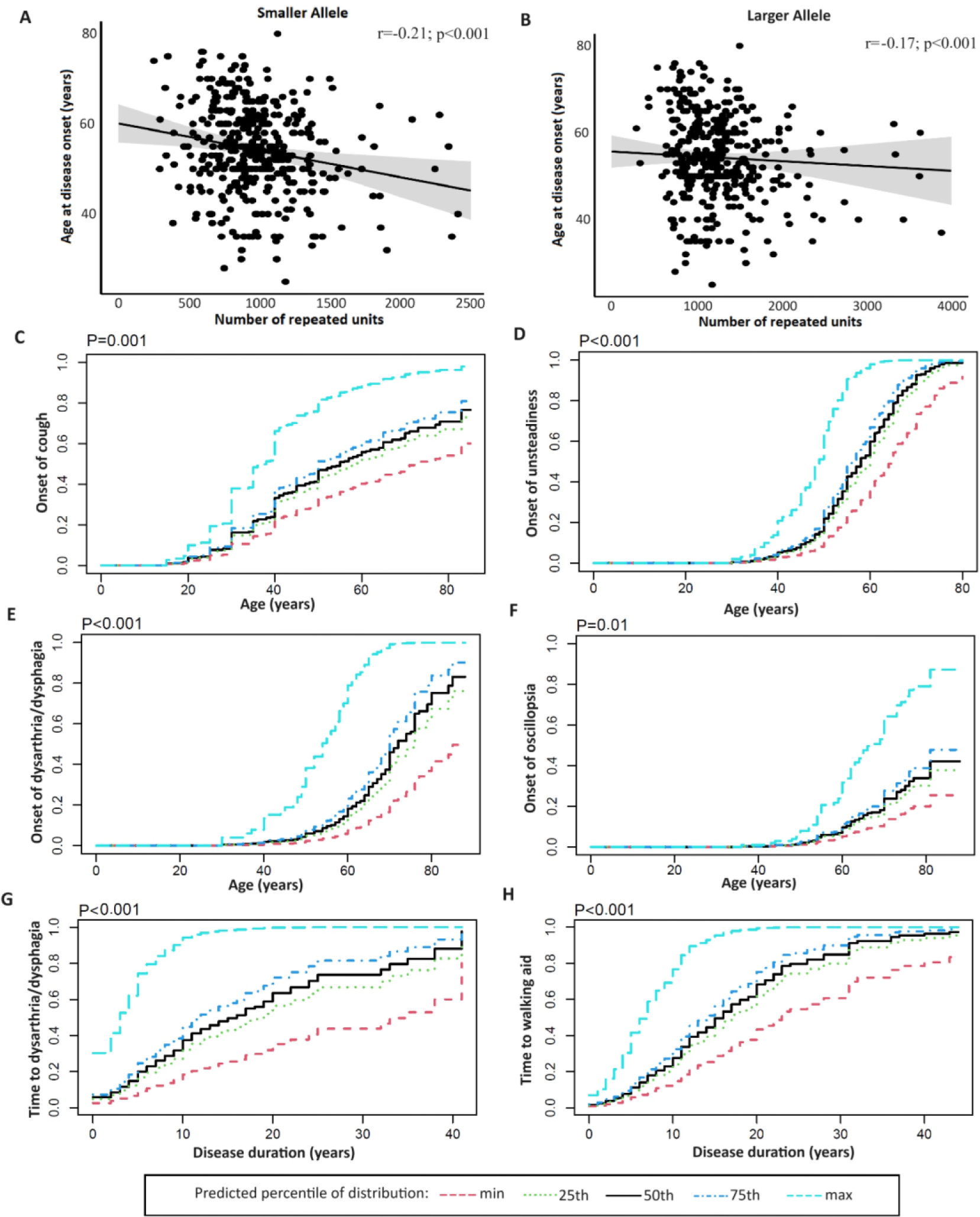
Relationship between repeat size and main symptoms of RFC1 disease. **(A-B)** The scatter plots illustrate the strength and the direction of the correlation between the age at neurological onset of the disease (Y axis) and the repeat size of the smaller or the larger allele (X axis). (**C-F)** the curves illustrate the predicted cumulative incidence function (CIF) for chronic cough, unsteadiness, dysarthria/dysphagia, and oscillopsia plotted against age at onset. (**G-H)** predicted CIF for dysarthria/dysphagia and need of walking aid plotted against disease duration. Curves are stratified for values of smaller allele repeat size equal to the minimum value, 25th, 50th,75th percentiles and maximum value of distribution.

A Fine-Gray model with robust cluster standard errors, adjusted for competing risk of death and corrected for gender, was adopted to analyse the effect of repeat size of smaller and larger allele on age at onset of main disease symptoms. Coefficients were calculated for 1000-repeat units increase.

The repeat size of the smaller and larger allele resulted to be significant predictors of age at onset of unsteadiness (*HR*=2.68, *p*<0.001 for the smaller allele; *HR*=1.64, *p*<0.001 for the larger allele) and at onset of dysarthria and/or dysphagia (*HR*=4.01, *p*<0.001 for the smaller allele; *HR*=1.93, *p*<0.001 for the larger allele). The repeat size of the smaller allele also correlated with the onset of cough (*HR*=1.95, *p*<0.001), whereas the larger allele showed a borderline association with the onset of sensory symptoms (*HR*=1.33, *p*=0.009 with adjusted threshold of significance *p*=0.01) (**figure 2C-2F and supplementary table 2s**). Patients carrying larger expansions had an increased risk to develop disabling symptoms earlier in disease course compared to individuals with smaller expansions (smaller allele: *HR*=3.40, *p*<0.001 for dysarthria/dysphagia and *HR*=2.78, *p*<0.001 for walking aids; larger allele: *HR*=1.71, *p*=0.002 for dysarthria/dysphagia and *HR*=1.60, *p*<0.001 for walking aids) (**figures 2G-2H**). However, age at disease onset was an independent predictors of disease course, with a later disease onset being associated with a shorter time to the onset of these symptoms (**supplementary table 3s).**

Median disease duration at onset of dysarthria/dysphagia and at need for walking aids was significantly shorter in patients with repeat size of the smaller allele above the 75^th^ percentile (14.5 years for dysarthria/dysphagia, 15.1 years for walking aids) compared to patients with a repeat size below the 25^th^ percentile (21.5 years for dysarthria/dysphagia, 19.2 years for walking aids; p<0.001).

### Repeat expansion size influences disease phenotype

After correcting for sex, age at last examination, and age at disease onset, the mean size of both alleles was significantly higher in patients with complex neuropathy (smaller allele *RR*=1.30, *p*=0.003; larger allele *RR*=1.33, *p*=0.008) or CANVAS (smaller allele *RR*=1.34, *p*<0.001; larger allele *RR*=1.31, *p*=0.009) than in patients with isolated neuropathy (**supplementary table 4s)**. This difference was confirmed by pair-wise comparisons of repeat size between the three phenotypes (**figure 3)**. Conversely, we did not observe a significant difference in repeat size between patients with complex neuropathy and CANVAS phenotype (*p*=0.83 smaller allele; *p*=0.97 larger allele).

**Figure 3.**
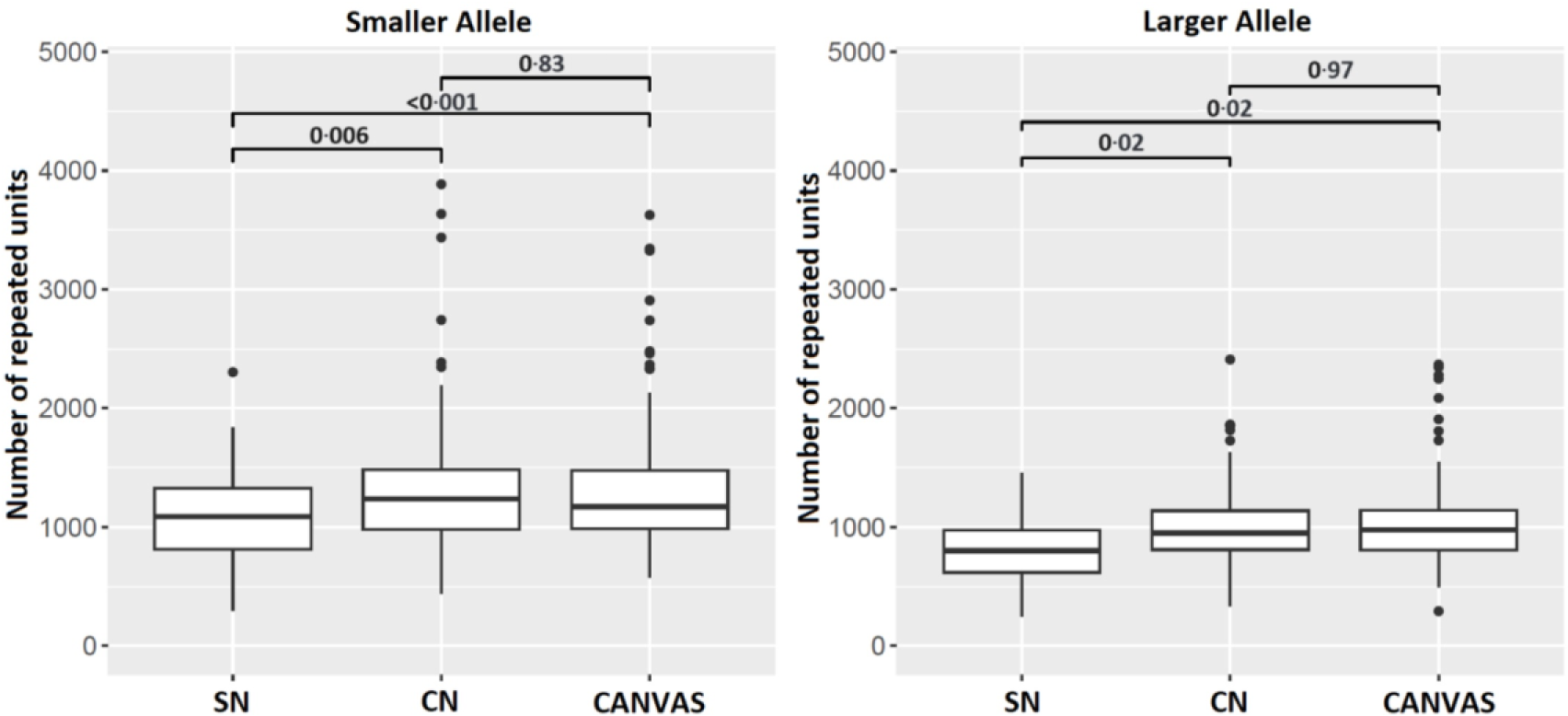
Relationship between phenotype and repeat length. The box plots compare the repeat size for the smaller (left panel) and larger (right panel) alleles in patients with different phenotypes at last examination. P-values were calculated adopting Tukey’s correction. SN= sensory neuropathy; CN= complex neuropathy.

### Repeat expansion size correlates with the degree of cerebellar atrophy

We next tested the association between the repeat length and the degree of cerebellar vermis atrophy in an internal cohort of 32 brain MRI performed at the National Hospital for Neurology and Neurosurgery, London (UK). After adjusting for age at MRI, disease duration and total intracranic volume, we observed a significant association (Bonferroni-adjusted significance level *α* = 0.017) between the repeat size of the smaller allele and the volume of cerebellar vermis lobules I-V (*β*=-1.06, *p*<0.001) and lobules VI-VII (*β*=-0.34, *p*=0.005). Conversely, the volume of lobules VIII-X did not correlate with the repeat size (β=-0.44, *p*= 0.07). No significant association was observed between the repeat size of the larger allele and cerebellar volume (**supplementary table 5s**).

### Stability of RFC1 repeat expansion across generations and tissues

We evaluated 69 subjects (including 27 probands, 22 siblings, 18 offspring and 2 parents) from 27 families, for a total of 64 meiotic events. AAGGG repeat expansion appeared stable across generations (*r^2^*=0.95), with a median intra-familial variation of 25 repeats (*IQR*=-17/+45, *min max*=-250/+510), and less than 10% compared to the proband’s allele in 80% of meiosis (**figure 4A**). Expansion or contraction of the repeat across generations occurred with the same frequency. Next, we compared the *RFC1* repeat size from different brain areas and peripheral tissues including blood, muscles and/or fibroblasts, as available, from four patients carrying biallelic *RFC1* expansions. There was limited instability of the repeat across the tissues analysed, with a variation in size between -97 and +190 repeats (-5%/+7%) compared with the mean size (**figure 4B**). Furthermore, mean dispersion of the repeat length was ± 1.7% for vermis, ± 2% for cerebellar hemispheres and ± 2.7% for frontal cortex, as opposed to a dispersion of ± 36% in an individual carrying *C9orf72* expansion. Overall, there was evidence of limited somatic instability across affected and unaffected bulk tissues.

**Figure 4.**
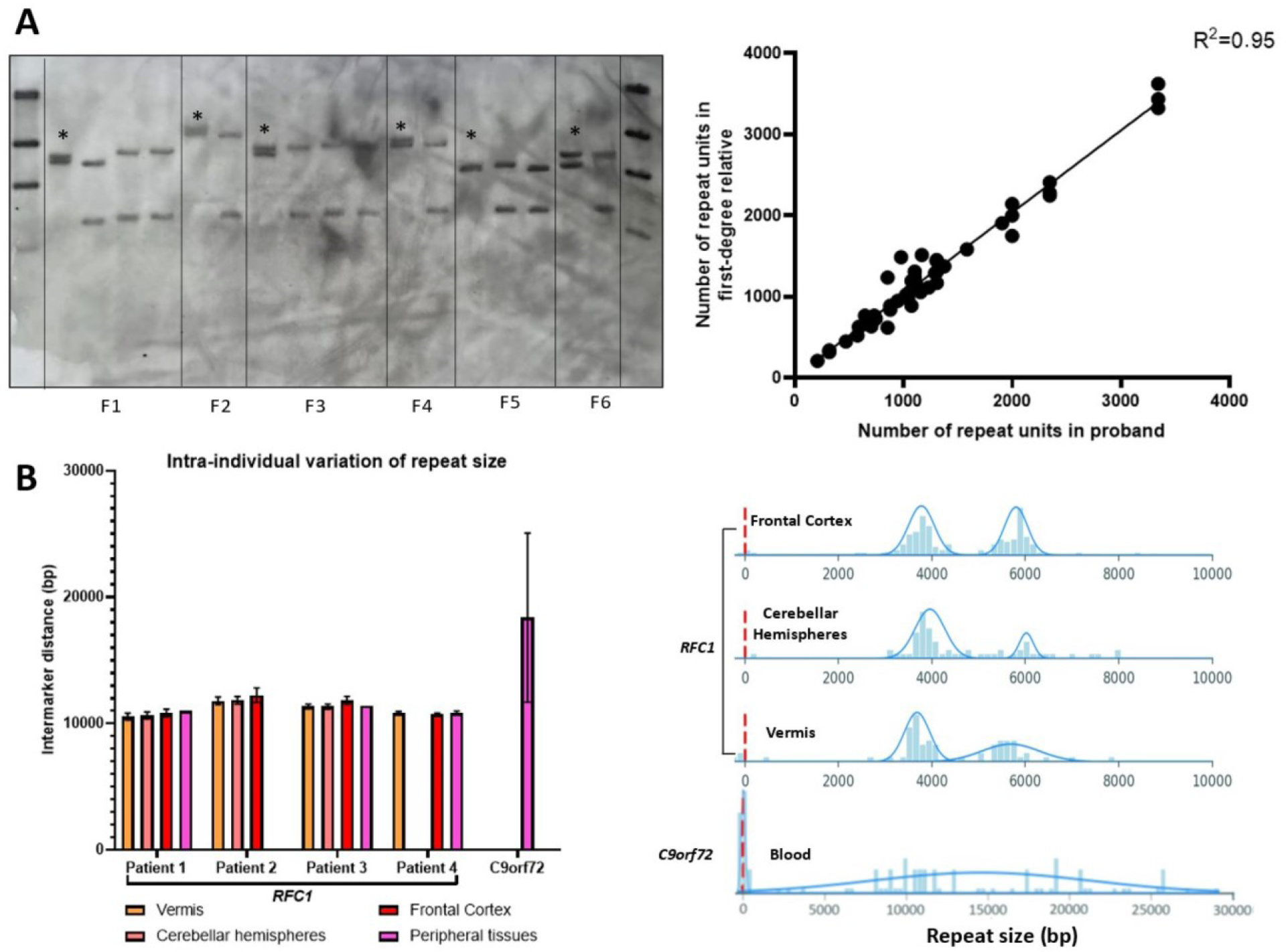
Limited meiotic and somatic instability of the AAGGG repeat expansion. **(A)** On the left, the picture illustrates a representative example of Southern Blotting including probands (asterisks) and unaffected relatives from six families (F1-F6). On the right, the correlation plot shows the relationship of the repeat size within members of the same family. Each dot corresponds to a meiotic event. **(B)** On the left, the bar chart shows the dispersion of the repeat size among different brain areas and peripheral tissues of four patients with RFC1 biallelic expansions and in one patient with C9orf72 expansion. Mean intermarker distance (expressed in base pairs) and SDs are shown. In patient 1 and patient 3, repeat size from peripheral tissue was measured by Southern blotting. On the right, distribution of DNA molecules measured by genome optical mapping (Bionano Genomics) in different tissues of a patient with a biallelic expansion in RFC1 and in blood-derived DNA of a patient with C9orf72 expansion. The size of DNA molecules mapping on RFC1 or C9orf72 locus is expressed in base pairs.

## Discussion

In this study we leveraged a large international cohort of genetically confirmed patients carrying biallelic *RFC1* expansions to assess the impact of the repeat expansion size on onset, clinical phenotype, and progression of *RFC1* disease.

Clinical data confirmed the existence of a spatial dissemination of the disease from an isolated sensory neuropathy with or without chronic cough to a complex neurodegenerative disease, mainly entailing clinically manifest cerebellar and vestibular involvement. Sensory neuropathy was present in all patients tested, confirming the central role of sensory involvement in *RFC1* disease. We showed that the repeat size in patients with isolated sensory neuropathy is smaller compared to patients with multisystem involvement and similar disease duration. Therefore, the repeat expansion acts as a modifier of the disease phenotype, probably due to a higher susceptibility of sensory neurons to the AAGGG repeat expansion, even of small size, compared to other tissues.

We observed a significant influence of the repeat length on the age of neurological onset. The association could be better appreciated when looking at well-defined clinical symptoms, like the onset of dysarthria and dysphagia, which tend to appear later in the disease course. Indeed, early sensory symptoms, imbalance and chronic cough may be initially mild and progress very slowly over time, so that patients struggle to date back the exact onset of the disease and often tend to date the first symptoms to few years before seeking neurologic attention.

Most importantly, we have demonstrated a direct impact of the repeat size on the disease severity and progression. Patients carrying larger expansions had a less favourable prognosis, with over 3-fold increased hazard of developing dysarthria or dysphagia and over 2-fold increased hazard of losing independent walking per 1000-units increase in repeat size. An older age at disease onset was associated with a faster progression. This has been observed in other neurodegenerative disorders, including sporadic and familiar (C9orf72) ALS/ALS-FTD^50–53^. It has been postulated that the faster progression observed in late-onset ALS cases might reflect the reduced neuronal reserve at baseline in older patients^51^. This hypothesis may also apply to *RFC1* disease. Alternatively, the shorter interval between disease onset and reaching disability milestones in cases with late onset may simply reflect a delayed diagnosis in individuals where early neurological symptoms, including sensory symptoms or mild unsteadiness, were overlooked or absent.

Importantly, we also showed that the repeat size of the smaller allele correlates with the degree of cerebellar vermis atrophy. The correlation is significant for lobules I-V and lobules VI-VII, in keeping with the selective atrophy of these lobules reported in previous neuroimaging and neuropathological studies^54,55^.

However, the degree of correlation might be influenced by the small sample size and by possible partial volume artifacts in 2D acquisitions. Prospective studies with larger cohorts and homogenous volumetric acquisitions are warranted to confirm these findings.

Importantly, the repeat size explained only up to 6% of the variability in age of neurological disease onset, suggesting that additional environmental or genetic modifiers at the repeat locus or in distant genes may be at play. In particular, the study was not designed to address the role of repeat interruptions, as Southern Blotting only provides information about the repeat size.

The study also demonstrated that the pathogenic AAGGG repeat has limited germline and somatic instability. Unlike most short tandem repeats, including the CAG repeat in Huntington disease and other polyglutamine diseases, CTG in myotonic dystrophy type 1, CGG in Fragile X syndrome-FXTAS, and CCGGGG in C9orf72, in which a significant instability of the expanded repeat was demonstrated,^8,48,56–59^ the *RFC1* AAGGG repeat appears stable across generations, with a repeat size variation, including contraction and further expansion, mostly unchanged or limited to 10% of repeat size.

To this regard, it is interesting to note that the size of the two alleles was not independent in the population tested and one third of cases had biallelic expansions of identical or highly similar size. We hypothesize that this observation could be caused by a geographic distribution of expanded alleles of a similar size, which is maintained over time and across generations, and we speculate that the stability of the *RFC1* repeat observed in single families may extend to broader areas and regions, inhabited by distantly related individuals.

To gain insight into the mechanisms underlying the tissue-specific involvement of *RFC1* disease, we tested whether the AAGGG repeat may undergo a further somatic expansion in the affected cerebellum. Although a variation of the repeat size at single cell level cannot be excluded, the data obtained from bulk tissue does not support the existence of significant instability of the repeat size as a determinant of the selective involvement of specific regions and neuronal populations in *RFC1* disease.

Finally, the relative stronger effect of the size of the smaller allele may suggest the existence of an underlying loss-of-function mechanism in *RFC1* disease, with progressive decrease of the residual *RFC1* function sustained by the allele carrying the smaller of the two expansions. This hypothesis is also supported by the recent identification of truncating variants in *RFC1* in compound heterozygous state with an expansion on the second allele, leading to typical, if not more severe, CANVAS phenotype.^32,33^ These observations are particularly relevant to the understanding of the disease-causing mechanism of *RFC1* disease since, despite the recessive mode of inheritance, the expression of *RFC1* transcript and protein seems unchanged.^23^ Notably, truncating variants in the coding region and a prominent effect of the smaller allele have both been previously observed in Friedreich’s ataxia, a recessive disorder caused by biallelic GAA expansion in *FXN*, and for which, as opposed to *RFC1* disease, a reduced expression and loss of function of the repeat containing gene has been clearly shown.^60^

The main limits of this work are related to its retrospective nature. In particular, the milder influence of the repeat size on some clinical features might be partly explained by the difficulty of patients in dating back the onset of their earliest symptoms, as well as by a lack of homogeneity in the clinical examinations and investigations performed in different centres.

In conclusion, the study contributed to better define the genotype-phenotype spectrum of *RFC1* disease and highlighted the key role of the repeat size as disease modifier. Larger expansions, in particular of the smaller allele, are associated with an earlier onset, a more complex phenotype, and a more aggressive disease progression. Estimating the size of the repeat expansion by Southern Blotting or alternative methods which became only recently available (e.g., optical genome mapping) is important not only to confirm the presence of biallelic expansions, but also to identify patients with higher risk to develop more complex and disabling phenotypes after a shorter disease duration and to better inform them and their families on prognosis. This may also impact the future design of trials as it will be key that patients in placebo/active drug groups will have a comparable distribution of repeat sizes.

## Data availability

With publication, de-identified data collected for the study, including individual participant data and a data dictionary defining each field in the set, can be made available to interested parties on reasonable request, and if in line with privacy regulations. Data can be requested at least 18 months after publication of this manuscript by sending an e-mail to the corresponding author.

## Data Availability

All data produced in the present study are available upon reasonable request to the authors

## Acknowledgements

We thank the patients and relatives who participated in this study.

## Funding

This work was supported by Medical Research Council (MR/T001712/1), Fondazione Cariplo (grant n. 2019-1836), the Inherited Neuropathy Consortium, and Fondazione Regionale per la Ricerca Biomedica (Regione Lombardia, project ID 1751723). R. Currò was supported by the European Academy of Neurology (EAN) Research Fellowship 2021. H. Houlden and M.M. Reilly thank the MRC, the Wellcome Trust, the MDA, MD UK, Ataxia UK, The MSA Trust, the Rosetrees Trust and the NIHR UCLH BRC for grant support. F. Taroni thanks the Fondazione Regionale per la Ricerca Biomedica (CP 20/2018 (Care4NeuroRare) and the Italian Ministry of Health (RC) for grant support. D. Pareyson thanks the Italian Ministry of Health (RRC) for grant support. F.M. Santorelli thanks Ricerca Corrente 2022 Ministero della Salute 5X1000 for grant support. M. Synofzik thanks the Deutsche Forschungsgemeinschaft (DFG, German Research Foundation) and the European Joint Programme on Rare Diseases for grant support. P.F. Chinnery the Medical Research Council Mitochondrial Biology Unit, the Medical Research Council (MRC) International Centre for Genomic Medicine in Neuromuscular Disease, the Leverhulme Trust (RPG-2018-408), the Medical Research Council, the Alzheimer’s Society Project, and the NIHR Cambridge Biomedical Research for grant support.

## Competing interests

The authors report no competing interests.

## Appendix 1 RFC1 repeat expansion study group

Inés Albájar, Catherine Ashton, Nick Beauchamp, Sarah J Beecroft, ^,^Emilia Bellone, Josè Berciano, Petya Bogdanova-Mihaylova, ^,^Barbara Borroni, Bernard Brais, Enrico Bugiardini, Catarina Campos, Aisling Carr, Liam Carroll, Francesca Castellani, Tiziana Cavallaro, Patrick F. Chinnery, Silvia Colnaghi, ^,^Giuseppe Cosentino, Joana Damasio, Soma Das, Grazia Devigili, Daniela Di Bella, David Dick, Alexandra Durr, Amar El-Saddig, Jennifer Faber, Moreno Ferrarini, Massimiliano Filosto, Geraint Fuller, Salvatore Gallone, ^,^Chiara Gemelli, Marina Grandis, John Hardy, Channa Hewamadduma, Rita Horvath, Vincent Huin, Daniele Imperiale, Pablo Iruzubieta, Diego Kaski, Andrew King, Thomas Klockgether, Müge Koç, ^,^Kishore R Kumar, Thierry Kuntzer, Nigel Laing, Matilde Laurà, Timothy Lavin, Peter Nigel Leigh, Lea Leonardis, Michael P Lunn, Stefania Magri, Francesca Magrinelli, Maria João Malaquias, Michelangelo Mancuso, Hadi Manji, ^,^Sara Massucco, John McConville, Renato P. Munhoz, ^,^Sara Nagy, Alain Ndayisaba, Andrea Hilary Nemeth, Luiz Eduardo Novis, Johanna Palmio, Elena Pegoraro, David Pellerin, Benedetta Perrone, Chiara Pisciotta, James Polke, Malcolm Proudfoot, Laura Orsi, Aleksandar Radunovic, Nilo Riva, Aiko Robert, ^,^Riccardo Ronco, Elena Rossini, Alex M Rossor, Irmak Şahbaz, Qais Sa’di, Ettore Salsano, Alessandro Salvalaggio, Lucio Santoro, Elisa Sarto, Andrew Schaefer, ^,^Angelo Schenone, Carolin Scriba, Joseph Shaw, Gabriella Silvestri, James Stevens, Michael Strupp, Charlotte J Sumner, Agnieszka Szymura, Matteo Tagliapietra, ^,^Cristina Tassorelli, Alessandra Tessa, Marie Theaudin, Pedro Tomaselli, Stefano Tozza, Arianna Tucci, Enza Maria Valente, Maurizio Versino, ^,^Richard A Walsh, Nick W Wood, Way Yan Yau, Stephan Zuchner

## Supplementary materials

**Table 1s.**
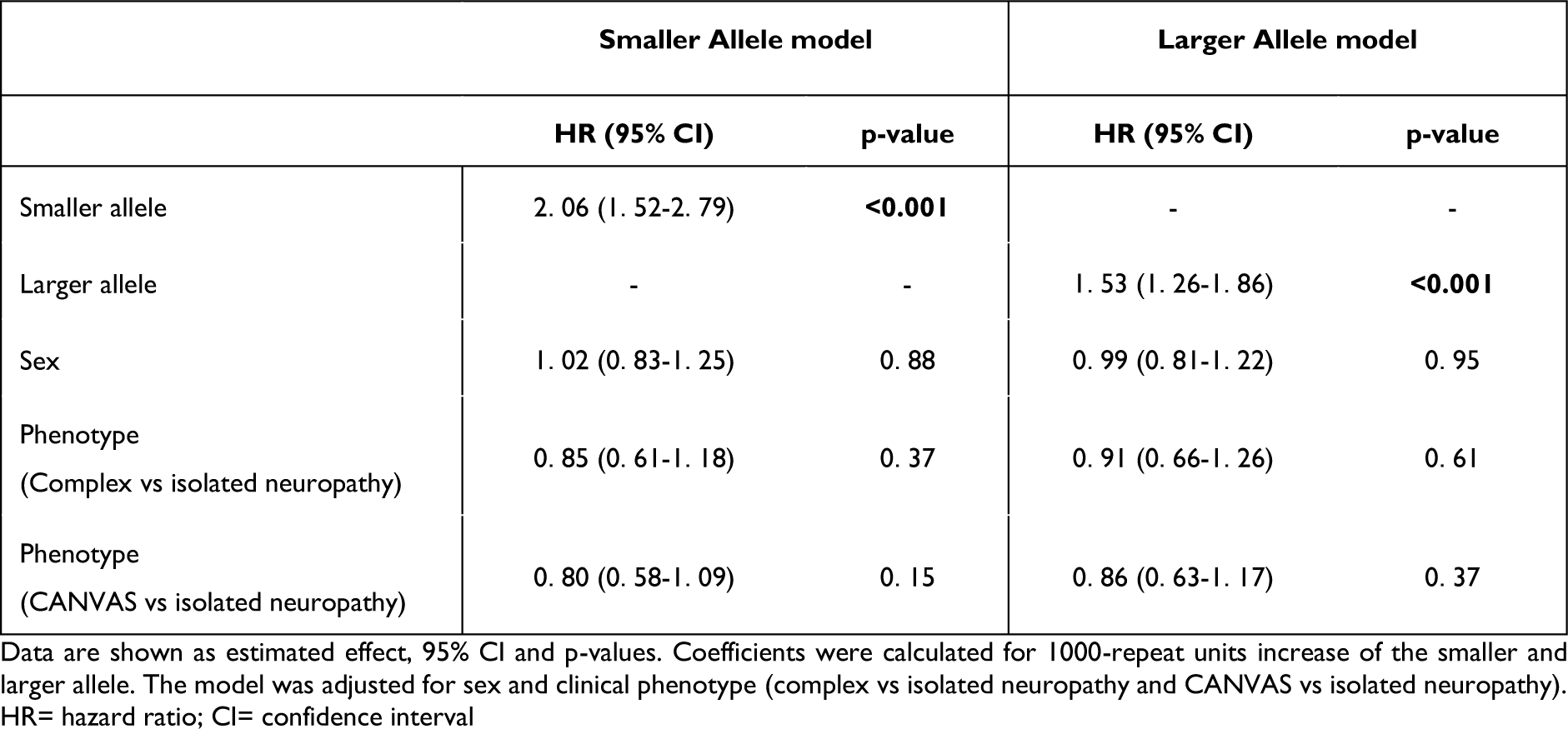
Cox regression analysis for RFC1 repeat size and age at neurological onset.

**Table 2s.**
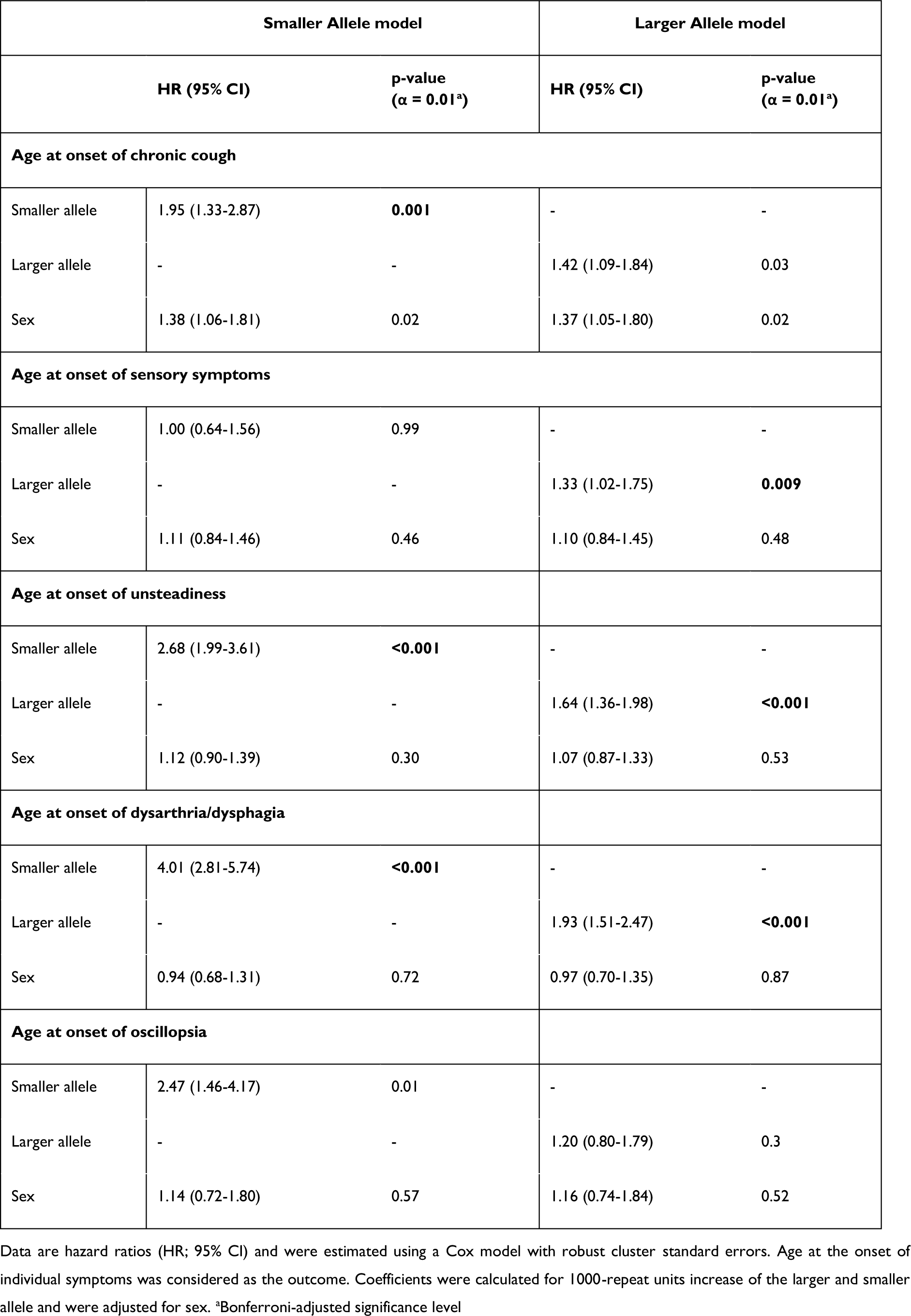
Cox regression analysis for RFC1 repeat size and age at onset of main symptoms.

**Table 3s.**
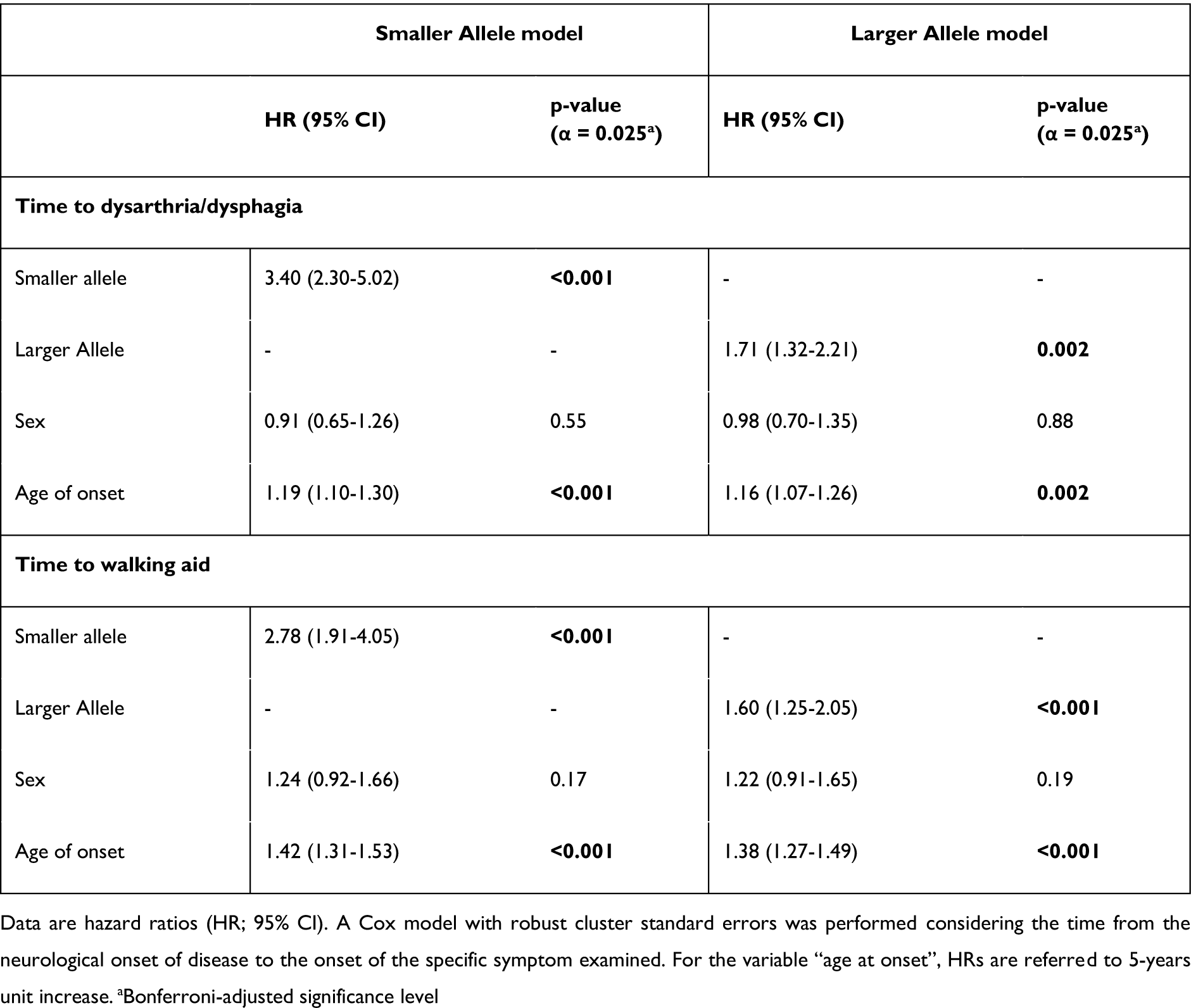
Cox regression analysis for RFC1 repeat size and time to disabling symptoms.

**Table 4s.**
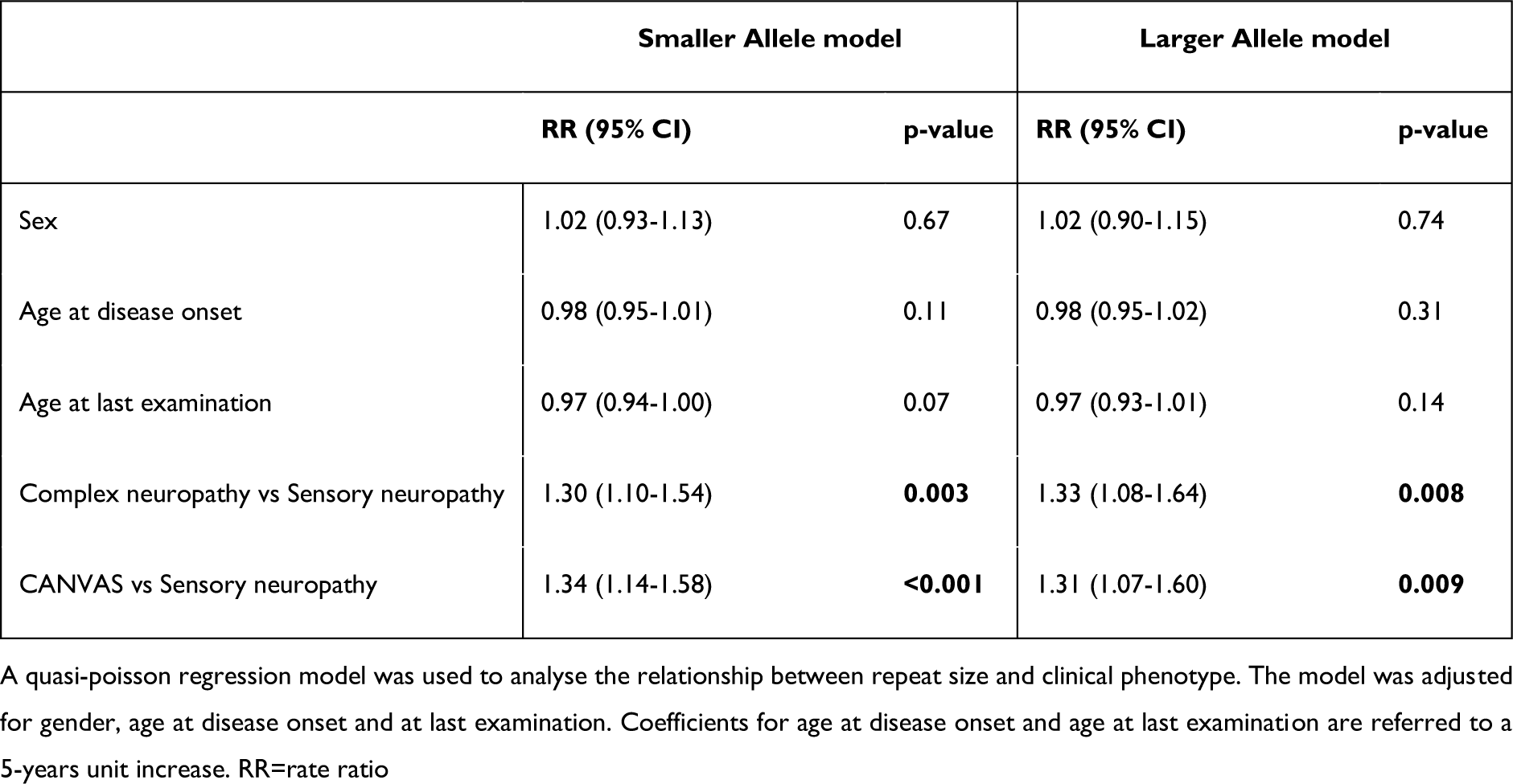
Quasi-poisson regression for RFC1 repeat size and disease phenotype.

**Table 5s.**
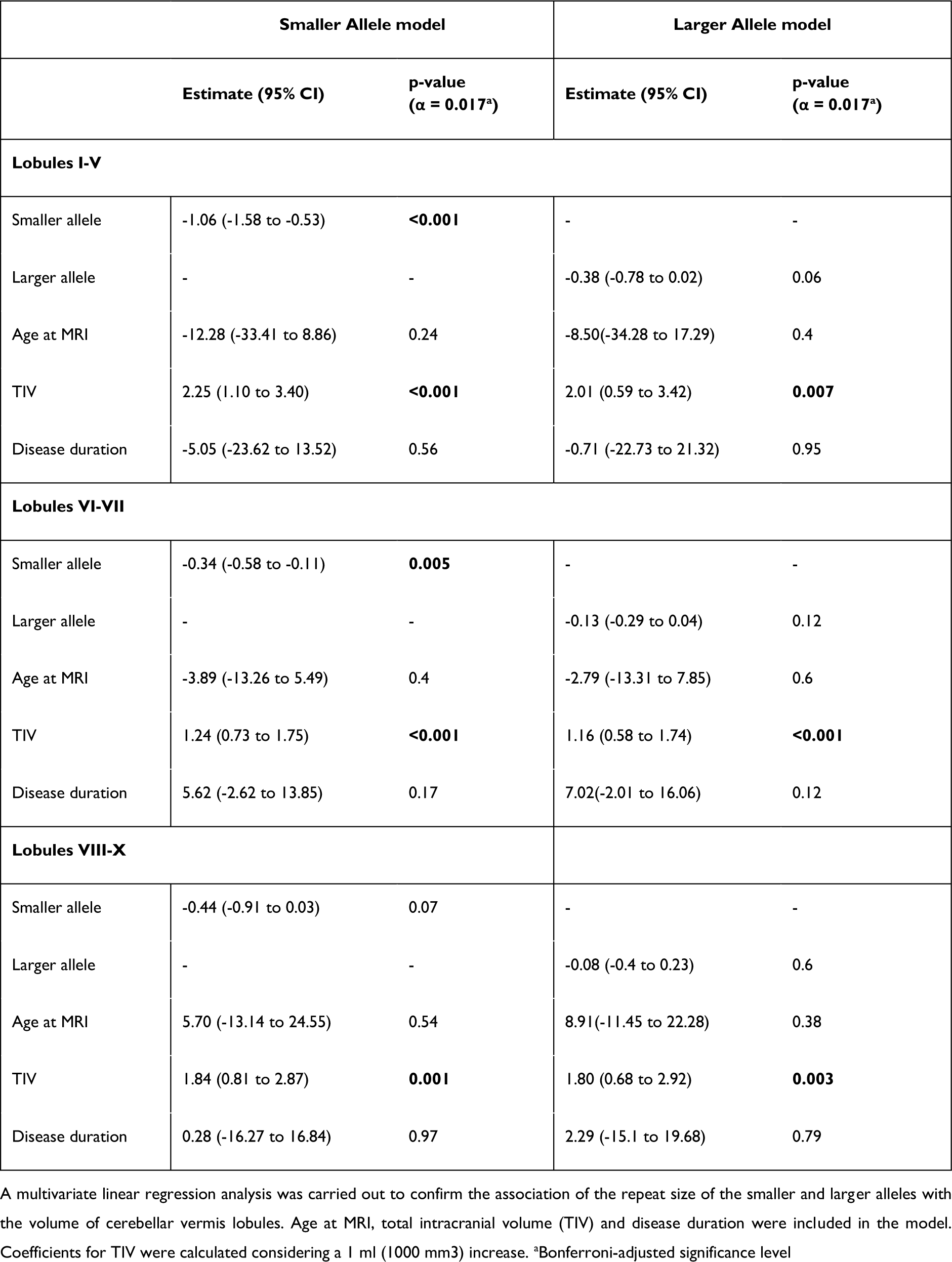
Multivariate linear regression analysis for RFC1 repeat expansion size and cerebellar vermis volume.

## Notes

### Competing Interest Statement

The authors have declared no competing interest.

### Author Declarations

Northeast-Newcastle & North Tyneside 1 Research Ethics Committee (22/NE/0080) gave ethical approval for this work.

