## Supplementary tables for "Role of the repeat expansion size in predicting age of onset and severity in RFC1 disease"

**Table 1s Cox regression analysis for RFC1 repeat size and age at neurological onset**

|  | **Smaller Allele model** | | **Larger Allele model** | |
| --- | --- | --- | --- | --- |
|  | **HR (95% CI)** | **p-value** | **HR (95% CI)** | **p-value** |
| Smaller allele | 2.06 (1.52-2.79) | **<0.001** | - | - |
| Larger allele | - | - | 1.53 (1.26-1.86) | **<0.001** |
| Sex | 1.02 (0.83-1.25) | 0.88 | 0.99 (0.81-1.22) | 0.95 |
| Phenotype  (Complex vs isolated neuropathy) | 0.85 (0.61-1.18) | 0.37 | 0.91 (0.66-1.26) | 0.61 |
| Phenotype  (CANVAS vs isolated neuropathy) | 0.80 (0.58-1.09) | 0.15 | 0.86 (0.63-1.17) | 0.37 |

Data are shown as estimated effect, 95% CI and p-values. Coefficients were calculated for 1000-repeat units increase of the smaller and larger allele. The model was adjusted for sex and clinical phenotype (complex vs isolated neuropathy and CANVAS vs isolated neuropathy). HR= hazard ratio; CI= confidence interval

**Table 2s Cox regression analysis for RFC1 repeat size and age at onset of main symptoms**

|  | **Smaller Allele model** | | **Larger Allele model** | |
| --- | --- | --- | --- | --- |
|  | **HR (95% CI)** | **p-value (α = 0.01^a^)** | **HR (95% CI)** | **p-value (α = 0.01^a^)** |
| **Age at onset of chronic cough** | | |  |  |
| Smaller allele | 1.95 (1.33-2.87) | **0.001** | - | - |
| Larger allele | - | - | 1.42 (1.09-1.84) | 0.03 |
| Sex | 1.38 (1.06-1.81) | 0.02 | 1.37 (1.05-1.80) | 0.02 |
| **Age at onset of sensory symptoms** | | |  |  |
| Smaller allele | 1.00 (0.64-1.56) | 0.99 | - | - |
| Larger allele | - | - | 1.33 (1.02-1.75) | **0.009** |
| Sex | 1.11 (0.84-1.46) | 0.46 | 1.10 (0.84-1.45) | 0.48 |
| **Age at onset of unsteadiness** | | |  |  |
| Smaller allele | 2.68 (1.99-3.61) | **<0.001** | - | - |
| Larger allele | - | - | 1.64 (1.36-1.98) | **<0.001** |
| Sex | 1.12 (0.90-1.39) | 0.30 | 1.07 (0.87-1.33) | 0.53 |
| **Age at onset of dysarthria/dysphagia** | | |  |  |
| Smaller allele | 4.01 (2.81-5.74) | **<0.001** | - | - |
| Larger allele | - | - | 1.93 (1.51-2.47) | **<0.001** |
| Sex | 0.94 (0.68-1.31) | 0.72 | 0.97 (0.70-1.35) | 0.87 |
| **Age at onset of oscillopsia** | | |  |  |
| Smaller allele | 2.47 (1.46-4.17) | 0.01 | - | - |
| Larger allele | - | - | 1.20 (0.80-1.79) | 0.3 |
| Sex | 1.14 (0.72-1.80) | 0.57 | 1.16 (0.74-1.84) | 0.52 |

Data are hazard ratios (HR; 95% CI) and were estimated using a Cox model with robust cluster standard errors. Age at the onset of individual symptoms was considered as the outcome. Coefficients were calculated for 1000-repeat units increase of the larger and smaller allele and were adjusted for sex. ^a^Bonferroni-adjusted significance level

**Table 3s Cox regression analysis for RFC1 repeat size and time to disabling symptoms**

|  | **Smaller Allele model** | | **Larger Allele model** | |
| --- | --- | --- | --- | --- |
|  | **HR (95% CI)** | **p-value (α = 0.025^a^)** | **HR (95% CI)** | **p-value (α = 0.025^a^)** |
| **Time to dysarthria/dysphagia** | | |  |  |
| Smaller allele | 3.40 (2.30-5.02) | **<0.001** | - | - |
| Larger Allele | - | - | 1.71 (1.32-2.21) | **0.002** |
| Sex | 0.91 (0.65-1.26) | 0.55 | 0.98 (0.70-1.35) | 0.88 |
| Age of onset | 1.19 (1.10-1.30) | **<0.001** | 1.16 (1.07-1.26) | **0.002** |
| **Time to walking aid** | | |  |  |
| Smaller allele | 2.78 (1.91-4.05) | **<0.001** | - | - |
| Larger Allele | - | - | 1.60 (1.25-2.05) | **<0.001** |
| Sex | 1.24 (0.92-1.66) | 0.17 | 1.22 (0.91-1.65) | 0.19 |
| Age of onset | 1.42 (1.31-1.53) | **<0.001** | 1.38 (1.27-1.49) | **<0.001** |

Data are hazard ratios (HR; 95% CI). A Cox model with robust cluster standard errors was performed considering the time from the neurological onset of disease to the onset of the specific symptom examined. For the variable “age at onset”, HRs are referred to 5-years unit increase. ^a^Bonferroni-adjusted significance level

**Table 4s Quasi-poisson regression for RFC1 repeat size and disease phenotype**

|  | **Smaller Allele model** | | **Larger Allele model** | |
| --- | --- | --- | --- | --- |
|  | **RR (95% CI)** | **p-value** | **RR (95% CI)** | **p-value** |
| Sex | 1.02 (0.93-1.13) | 0.67 | 1.02 (0.90-1.15) | 0.74 |
| Age at disease onset | 0.98 (0.95-1.01) | 0.11 | 0.98 (0.95-1.02) | 0.31 |
| Age at last examination | 0.97 (0.94-1.00) | 0.07 | 0.97 (0.93-1.01) | 0.14 |
| Complex neuropathy vs Sensory neuropathy | 1.30 (1.10-1.54) | **0.003** | 1.33 (1.08-1.64) | **0.008** |
| CANVAS vs Sensory neuropathy | 1.34 (1.14-1.58) | **<0.001** | 1.31 (1.07-1.60) | **0.009** |

A quasi-poisson regression model was used to analyse the relationship between repeat size and clinical phenotype. The model was adjusted for gender, age at disease onset and at last examination. Coefficients for age at disease onset and age at last examination are referred to a 5-years unit increase. RR=rate ratio

**Table 5s Multivariate linear regression analysis for RFC1 repeat expansion size and cerebellar vermis volume**

|  | **Smaller Allele model** | | **Larger Allele model** | |
| --- | --- | --- | --- | --- |
|  | **Estimate (95% CI)** | **p-value (α = 0.017^a^)** | **Estimate (95% CI)** | **p-value (α = 0.017^a^)** |
| **Lobules I-V** | | |  |  |
| Smaller allele | -1.06 (-1.58 to -0.53) | **<0.001** | - | - |
| Larger allele | - | - | -0.38 (-0.78 to 0.02) | 0.06 |
| Age at MRI | -12.28 (-33.41 to 8.86) | 0.24 | -8.50(-34.28 to 17.29) | 0.4 |
| TIV | 2.25 (1.10 to 3.40) | **<0.001** | 2.01 (0.59 to 3.42) | **0.007** |
| Disease duration | -5.05 (-23.62 to 13.52) | 0.56 | -0.71 (-22.73 to 21.32) | 0.95 |
| **Lobules VI-VII** | | |  |  |
| Smaller allele | -0.34 (-0.58 to -0.11) | **0.005** | - | - |
| Larger allele | - | - | -0.13 (-0.29 to 0.04) | 0.12 |
| Age at MRI | -3.89 (-13.26 to 5.49) | 0.4 | -2.79 (-13.31 to 7.85) | 0.6 |
| TIV | 1.24 (0.73 to 1.75) | **<0.001** | 1.16 (0.58 to 1.74) | **<0.001** |
| Disease duration | 5.62 (-2.62 to 13.85) | 0.17 | 7.02(-2.01 to 16.06) | 0.12 |
| **Lobules VIII-X** | | |  |  |
| Smaller allele | -0.44 (-0.91 to 0.03) | 0.07 | - | - |
| Larger allele | - | - | -0.08 (-0.4 to 0.23) | 0.6 |
| Age at MRI | 5.70 (-13.14 to 24.55) | 0.54 | 8.91(-11.45 to 22.28) | 0.38 |
| TIV | 1.84 (0.81 to 2.87) | **0.001** | 1.80 (0.68 to 2.92) | **0.003** |
| Disease duration | 0.28 (-16.27 to 16.84) | 0.97 | 2.29 (-15.1 to 19.68) | 0.79 |

A multivariate linear regression analysis was carried out to confirm the association of the repeat size of the smaller and larger alleles with the volume of cerebellar vermis lobules. Age at MRI, total intracranial volume (TIV) and disease duration were included in the model. Coefficients for TIV were calculated considering a 1 ml (1000 mm3) increase. ^a^Bonferroni-adjusted significance level
